# HUMAN PAPILLOMAVIRUS SEROTYPES IN ABNORMAL CERVICAL SMEARS IN CALABAR CROSS RIVER STATE A CROSS SECTIONAL STUDY

**DOI:** 10.1101/2022.05.17.22275089

**Authors:** Ocheze Chinwendu Orji, Edu Michael Eyong, Martins Anadozo Nnoli, Theophilus Ipeh Ugbem, Modupeola Samaila, Solomon Oladapo Rotimi, Ima-obong Asuquo Ekanem

## Abstract

**Background:** Human Papillomavirus (HPV) is sexually transmitted and constitutes the major cause of cancer of the cervix and could be detected using cervical smear screening test.

**Aim and objective:** To determine the frequency of high risk HPV serotypes (16,18,33,45)in abnormal cervical smears in women aged 18-65 years in Calabar, Cross River State using cytological method over a period of one year from 1^st^ March, 2017 to 28^th^ February, 2018 in Cross River State, Nigeria.

**Materials and method:** The study was a cross sectional study carried out in University of Calabar Teaching Hospital, Calabar on women aged 18-65 years who met the study inclusion criteria and were consecutively selected for conventional cervical screening test from the three provinces of Efik, Efut and Qua provinces. Females who were positive for squamous intraepithelial lesion after screening results were subjected to Human papillomavirus deoxyribonucleic acid (HPV DNA) testing.

**Results:** 304 women aged 18-65 years were recruited into the study. Of these only 30 had intraepithelial lesions. Low grade squamous lntraepithelial lesions (LSIL) predominated with 63.3%(19) as compared with High grade squamous lntraepithelial lesion of 36.6%(11). HPV DNA was observed in all the abnormal cytology subjected to HPV DNA test. The prevalence rate of high risk HPV in this study was 9.9 %. The commonest serotypes found among study participants were HPV 16,33,and 45 with prevalence of 4.7%, 100% and 61.9% respectively.

Among participants infected with HPV 33 and HPV 45, 13(61.9%) and 8(38.1%) had low grade squamous Intraepithelial lesion respectively while 8(38.1%) and 5(23.8%) had high grade squamous Intraepithelial lesion respectivelyIn this study,66.67%(14) out of 21 participants had HPV DNA co – infection.

**Conclusion:** The prevalent HPV serotypes were 16,33 and 45 from this study with serotypes 33 being the commonest. Advocacy for HPV vaccine for reproductive age females should be encouraged and this should be included with established cervical cancer screening programs in government hospitals.

## INTRODUCTION

Infectious agents mostly viruses contribute 13% of all cancer reports worldwide [1]infectious agents are essential factors to the world cancer burden especially in Africa [2].

A high infection-related cancer burden in sub-Saharan Africa was mainly cause by Human papilloma virus (HPV)[3]. HPVis among the commonest sexually transmissible infection (STI) globally[4].Cervical cancer is the fourth most common cancer found among women worldwide, with about 604 000 new cases and 342 000 deaths in 2020. Nearly 90% of the new cases and deaths globally in 2020 were seen in low- and middle-income countries [5].

Most cases of cervical cancer (more than 95%) is as a result of the human papillomavirus (HPV) infection[5].

There are more than 200 HPV types which have been known[6]. .HPVs are classified on the basis of their epidemiological relationship with cervical cancer into carcinogenic [high risk] non carcinogenic [low risk] subtypes. Out of these, 14 are classified as high-risk (HR) HPV types, they include types 16, 18, 31, 33, 35, 39, 45, 51, 52, 56, 58, 59, 68 and 73. [7], [8]. Although HR HPV types account for mostly all HPV-associated cancers, HPV 16 and 18 are the major causes of 70% of cases globally [9].

Cervical cancer starts from continuous infection by the high-risk human papilloma virus.(HPV) [10],[11]. Many studies confirmed that continuous infection with an oncogenic HPV type is the major risk factor for discovering a cervical intraepithelial neoplasia (CIN) that may start from CIN1 to CIN3 and then cancer [12], [13].

An essential step in cervical carcinogens is found in invasive cervical cancers encompasses HPV fusion into the host genome via clathrin-mediated endocytosis after binding to heparan sulfate proteoglycan (HSPG) and growth factor receptors (GFRs) on basal epithelial cells of the host following an injury [14], [15], [16], [17].The end result yield reproduction of viral genomes and their duplications.

The viral proteins E6 and E7 bind to tumour suppressor genes of the host predominately P53 and retinoblastoma genes respectively. [18],[19],[20],[21].

The net effect would be their degradation via proteasome pathway [18],[21].Final effect result in increase and uncoordinated proliferation of cells that acquire additional mutation which progress to carcinoma of cervix [21],[22].

Cervical cancer is an essential health and economic issue in Nigeria and globally. The well known cervical screening using Pap smear testing for diagnosis of abnormal cervical cells has greatly aided in early detection of the cervical cancer. Progress in HPV DNA cervical screening results has helped in the development of vaccines that prevent HPV infection.

## Materials and Methods

This study protocol was reviewed and approved by Health Research Ethics Committee, University of Calabar Teaching Hospital. Approval Protocol Number and National Health Research Ethics Committee Registration Number UCTH/HREC/33/512 and NHREC/07/10/2012 respectfully. Informed written consent was obtained from the participants after explaining to them the objectives of the study. All information collected on each individual were held in confidence. The Consecutive selection of women who met the study inclusion criteria for cervical screening from the three provinces of Qua, Efut and Efik Provinces in Calabar were recruited into the study after an awareness was created. The inclusion criteria were, females who were sexually active and aged 18-65years and females that had given birth more than 12 weeks, all residing more than ten years in Calabar, Cross River State. Females who did not consent, or were pregnant, or had hysterectomy or history of cervical pathology were excluded.

The sample size for the study was determined using the statistical formula for cross-sectional descriptive study. A total number of 304 women were selected by consecutive sampling method for the study.

All participants signed written informed consent formsbefore preceding to enrolment in the study and were given questionnaires after propercounsellingabout the objective and methodology of the study. Questionnaires were directed to data such as socio-demographic, reproductive, sexual histories, family histories, histories of biopsies and visual inspections of cervix.

The examiners ensured exact confidentiality of all participants’ information

The participants were discouraged of the following; use of vaginal cream and tablets, douching, involvingin sexual activity between 24 to 48 hours prior to the test.

The participants were then subjected to a complete pelvic examination Conventional cervical samples were obtained using sterile disposable Ayre’s spatulas. The smears collected were quickly fixed to slides before being transferred to the laboratory for processing. The fixed smears were stained using the Papanicolaou staining procedure The results of the smears were reported using modified Bethesda System 2014 version [23],[24].

The Participants that had epithelial cell abnormalities: those with High grade Squamous Intraepithelial Lesion (HSIL) and Low Grade Squamous Intraepithelial Lesions (LSIL) were called back ten days after the screening results came out, in order to retrieve another sample using DNA Swab for DNA studies.

The Participants that were Negative for Squamous Intraepithelial Lesions/Malignancy were not subjected to any further test rather they were advised on the need for routine pap smear examination based on their age as recommended by World Health Organization.

The samples that were collected from the participants that had LSIL and HSIL were suspended in a DNA Stabilizing Solutions, stored immediately in the Fridge at temperature of –80 degrees celsius later proceed for molecular analyses by means of DNA isolation techniques and polymerase chain reaction (PCR). using consensus primers targeted against the high risk HPV viruses.

The molecular study was done in Department of Biochemistry, Molecular Diagnostic laboratory, Covenant University, Ota, Ogun State. The extraction and purification of the viral HPV DNA was done using QiagenR deoxyribonucleic acid extraction kits while amplification of the extracted viral DNA was done using multiplex polymerase chain reaction guided by the manufacturers direction with the use of particular HPV primers for the high-risk types. The Multiplex PCR method was made by designing 4 type-specific primer sets that target conserved regions of the L1 gene of HPV which therefore allowed the amplification of strictly conserved regions of L1 gene in order to identify the HPV genotypes 16, 18, 33, 45,and using the human b-actin gene as internal control. Observation of amplified viral DNA was accomplished by agarose gel electrophoresis. All quantitative data were analyzed using Social Package for the Social Sciences version 23.0. Descriptive statistics were then computed for all relevant data. Chi Square Test to confirm relationship between categorized variables was used and Fisher’s Exact Test where appropriate.

## RESULTS -CYTOLOGY

The following are the photomicrographs of cytological representative results.

### Age characteristics of study participants

Table 1 below shows the age characteristics of study participants. A total of 304 women aged from 18 to 65 years participated in the study. The highest age group was 36-45 years with a proportion of 106(34.9%), followed by those aged 26-35 years with a proportion of 91(29.9%), then those in the 46-55 years age group with a proportion of 65(21.4%). The mean age of the study participants was 40.5 ± 10.2 years

**Table 1:**
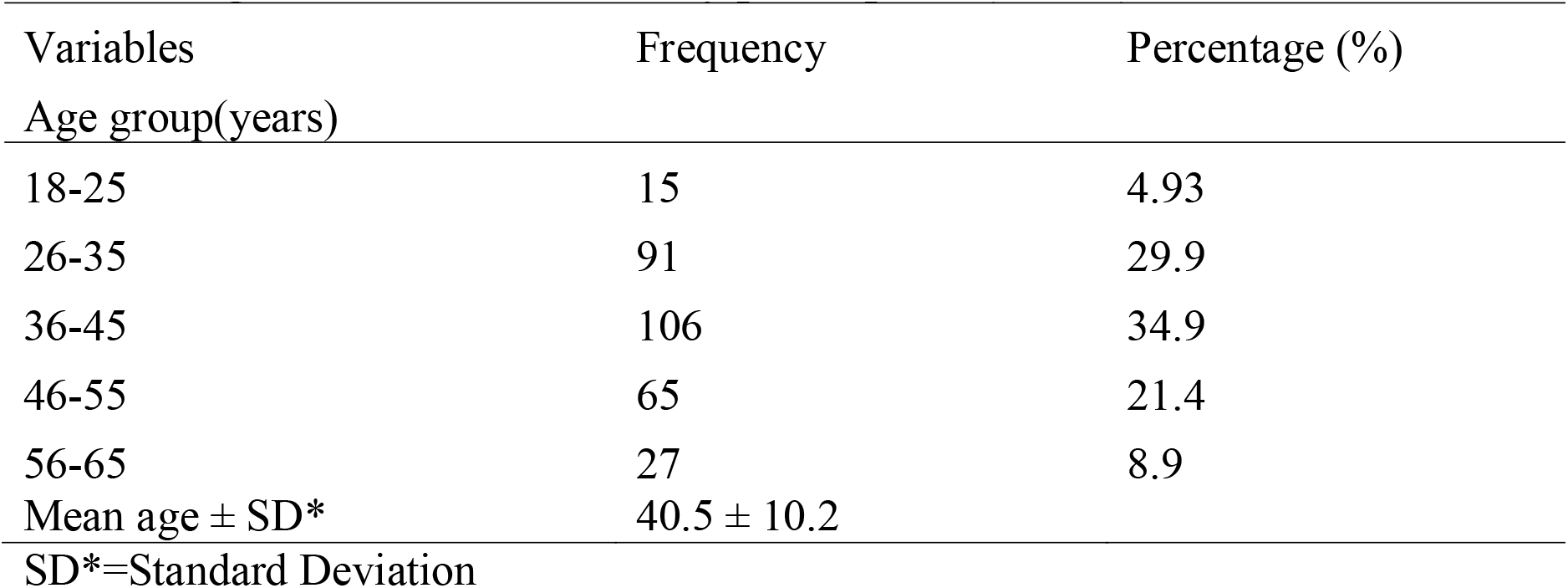
Age characteristics of study participants (N=304)

**Table 2:**
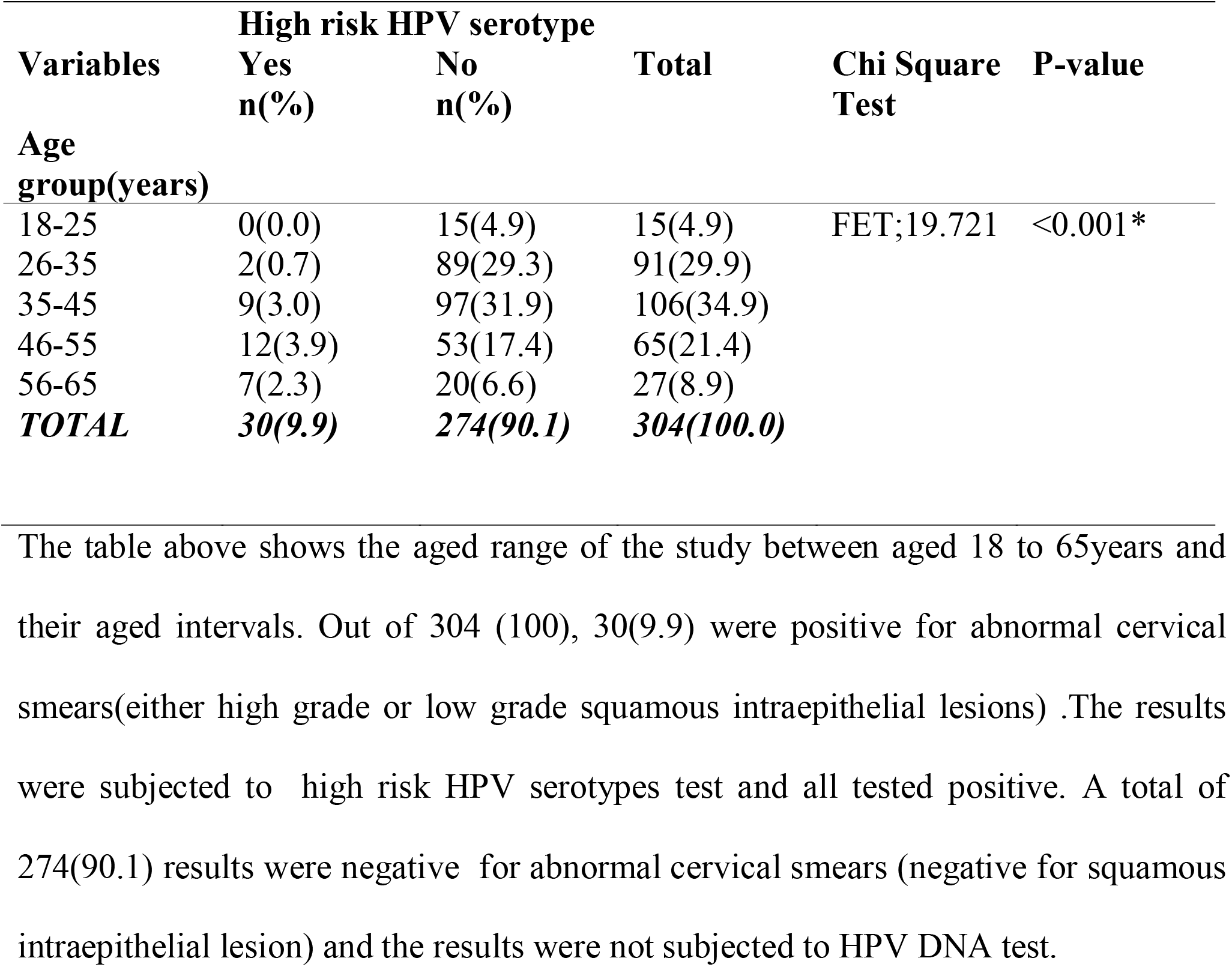
Association between age characteristics and high risk HPV serotype predominance among study participants.

| Variables | High risk HPV serotype |  | Total | Chi Square Test | P-value |
| --- | --- | --- | --- | --- | --- |
|  | Yes<br>n(%) | No<br>n(%) |  |  |  |
| **Age group(years)** |  |  |  |  |  |
| 18-25 | 0(0.0) | 15(4.9) | 15(4.9) | FET;19.721 | <0.001* |
| 26-35 | 2(0.7) | 89(29.3) | 91(29.9) |  |  |
| 35-45 | 9(3.0) | 97(31.9) | 106(34.9) |  |  |
| 46-55 | 12(3.9) | 53(17.4) | 65(21.4) |  |  |
| 56-65 | 7(2.3) | 20(6.6) | 27(8.9) |  |  |
| **TOTAL** | **30(9.9)** | **274(90.1)** | **304(100.0)** |  |  |

The table above shows the aged range of the study between aged 18 to 65years and their aged intervals. Out of 304 (100), 30(9.9) were positive for abnormal cervical smears(either high grade or low grade squamous intraepithelial lesions) .The results were subjected to high risk HPV serotypes test and all tested positive. A total of 274(90.1) results were negative for abnormal cervical smears (negative for squamous intraepithelial lesion) and the results were not subjected to HPV DNA test.

### High risk HPV positive women

Figure 2 below is a bar chart showing HPV serotype among correspondent with high risk women. The most common serotype found among study participants was HPV 33 found among 100% (21), followed by HPV 45 found among 61.9%(13)then HPV16 which was found among 4.7%(1) and HPV 18 was 0(none) HPV16 which was found among 4.7%(1) and HPV 18 was 0(none).

**FIGURE 1:**
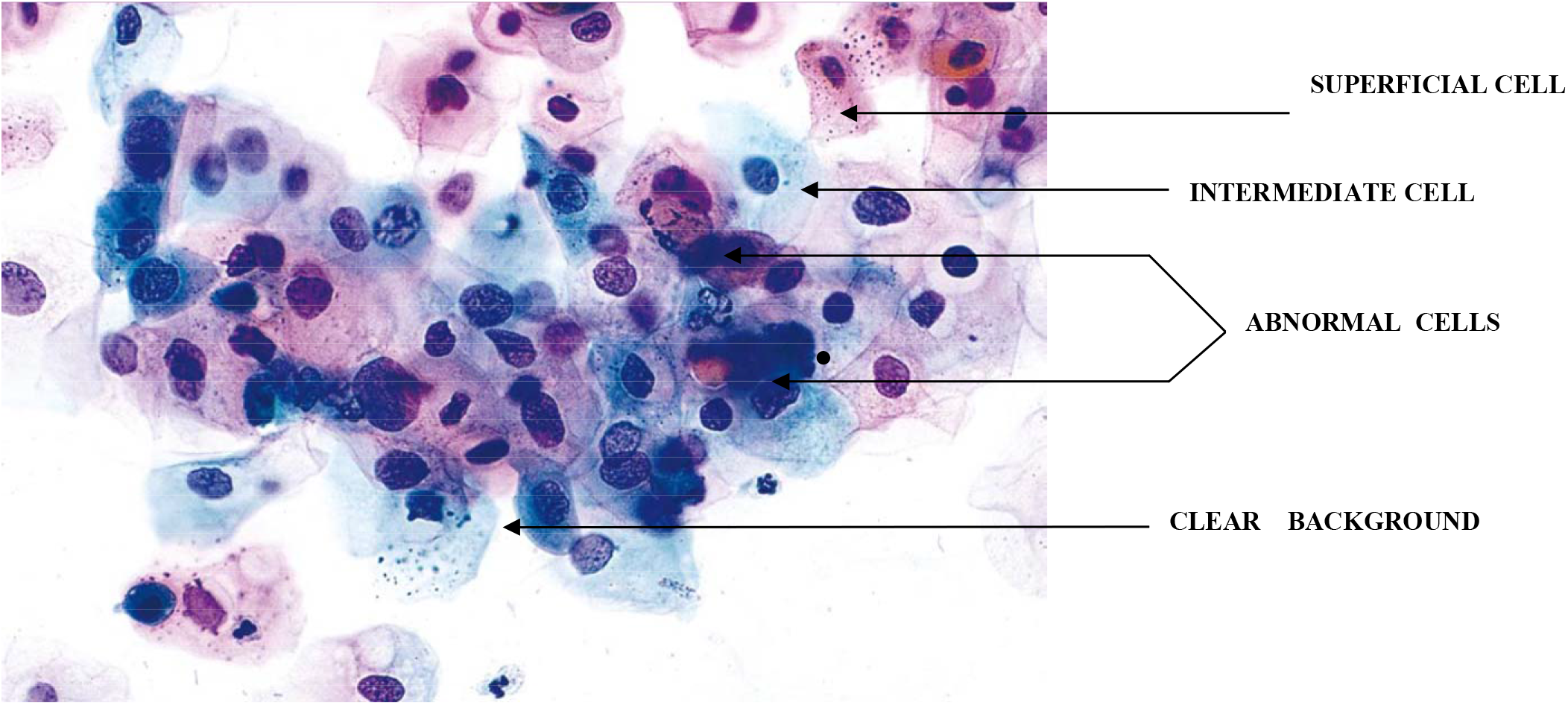
Low Grade Squamous Intraepithelial Lesion (papanicolaou stains X 40)

**FIGURE 2:**
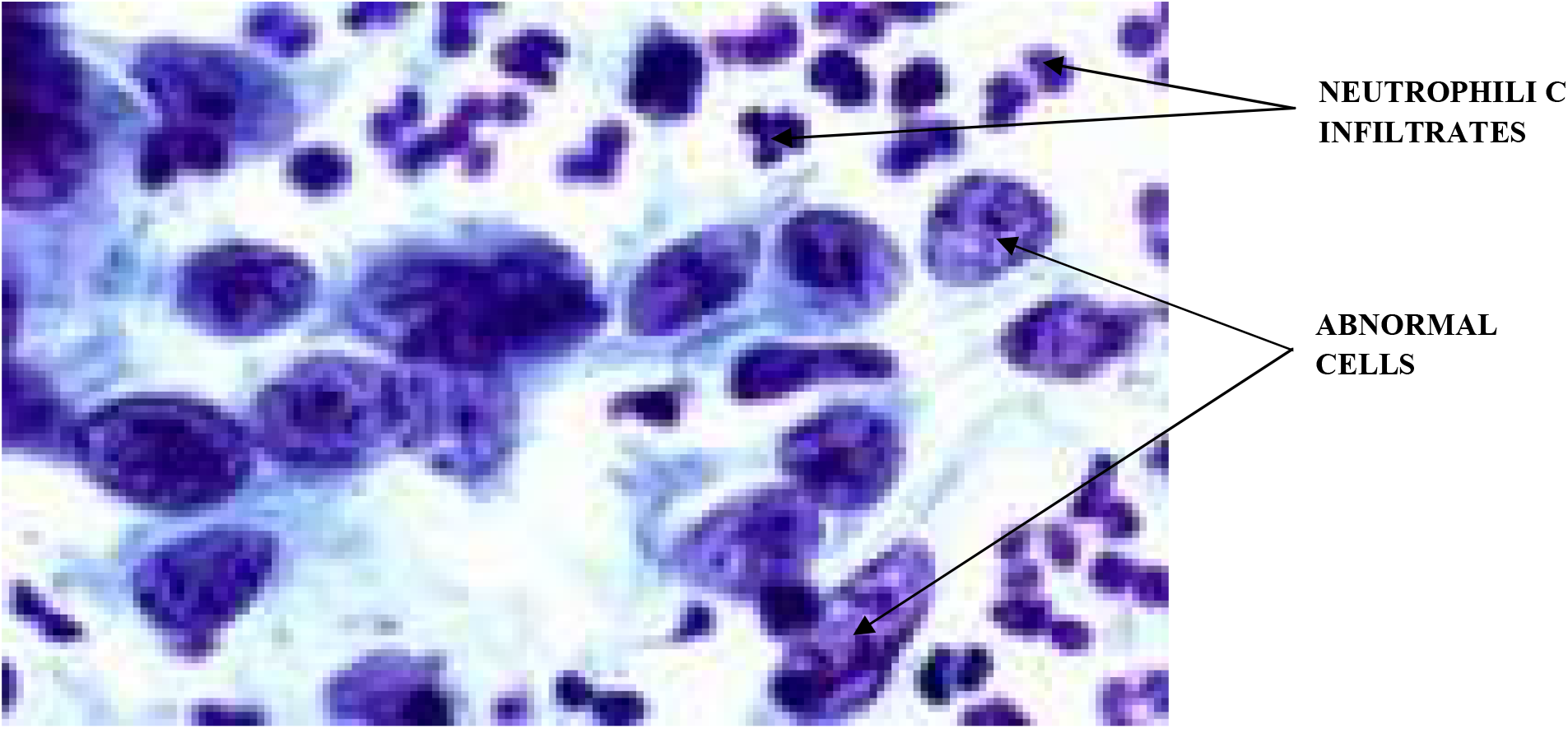
High Grade Squamous Intraepithelial Lesion (Papanicolaou stains X40)

**FIGURE 3:**
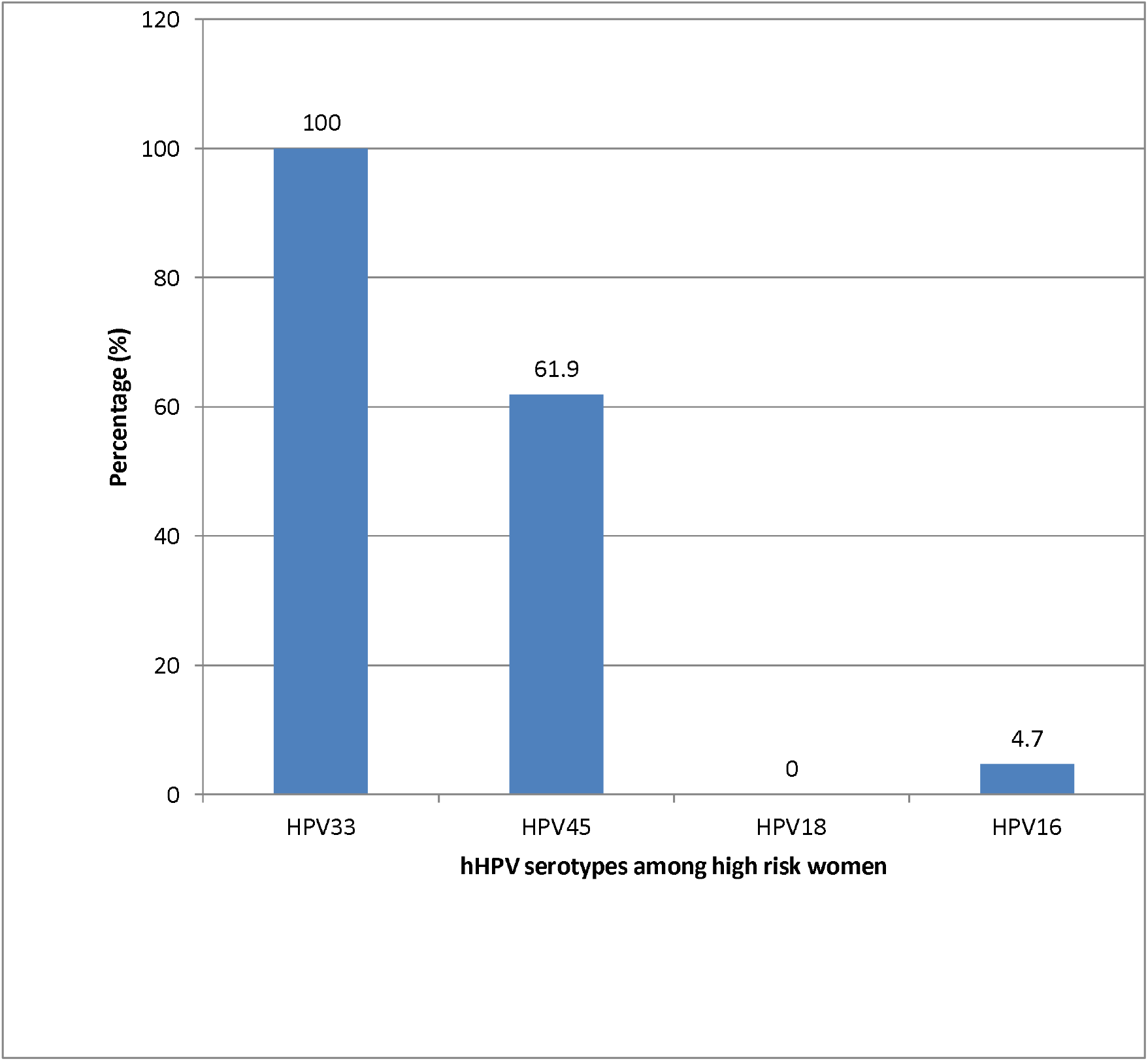
HPV serotype among high risk HPV positive women.

**FIGURE 4:**
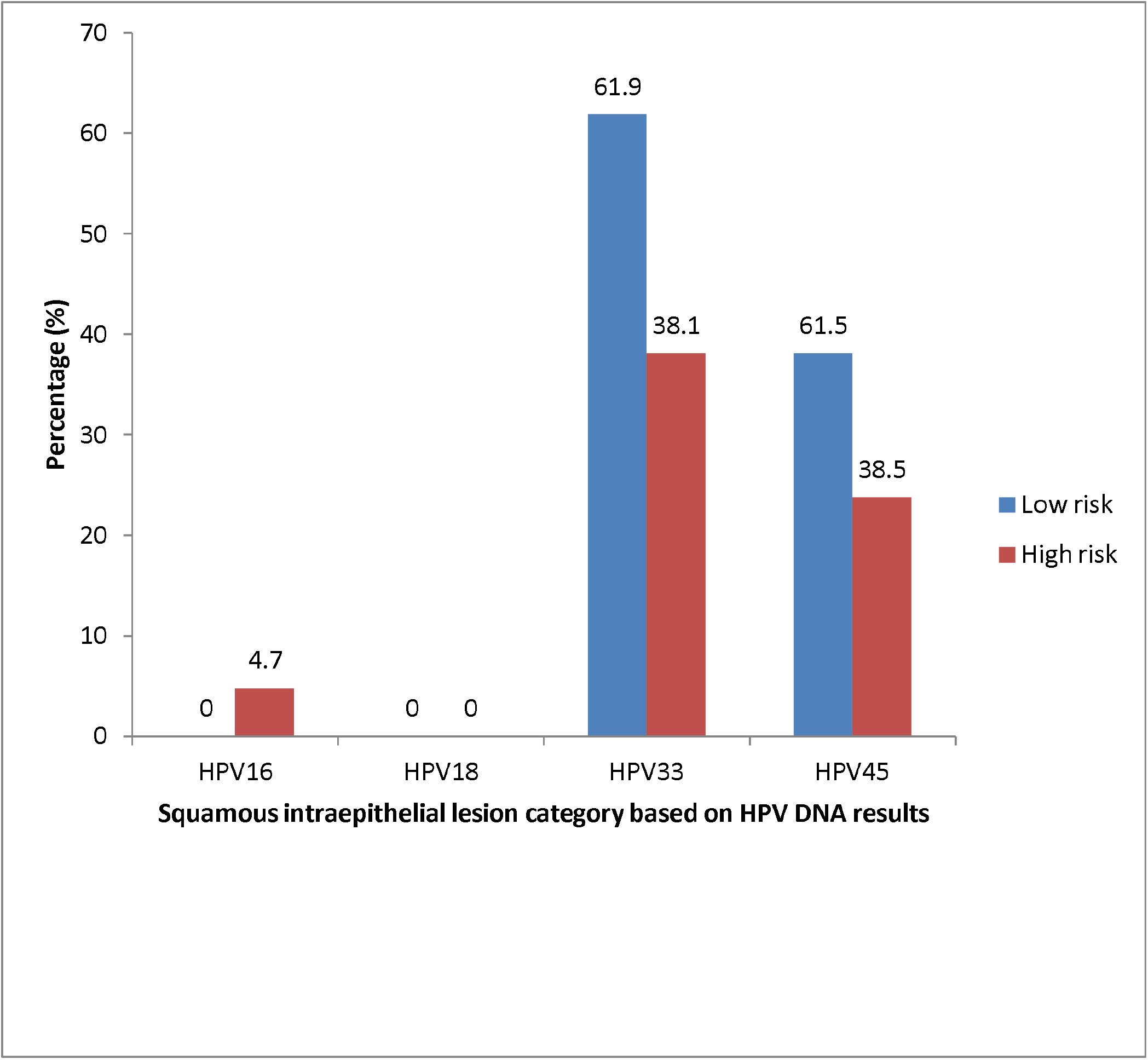
Squamous intraepithelial lesion category based on HPV DNA results as described by the table above.

**FIGURE 5:**
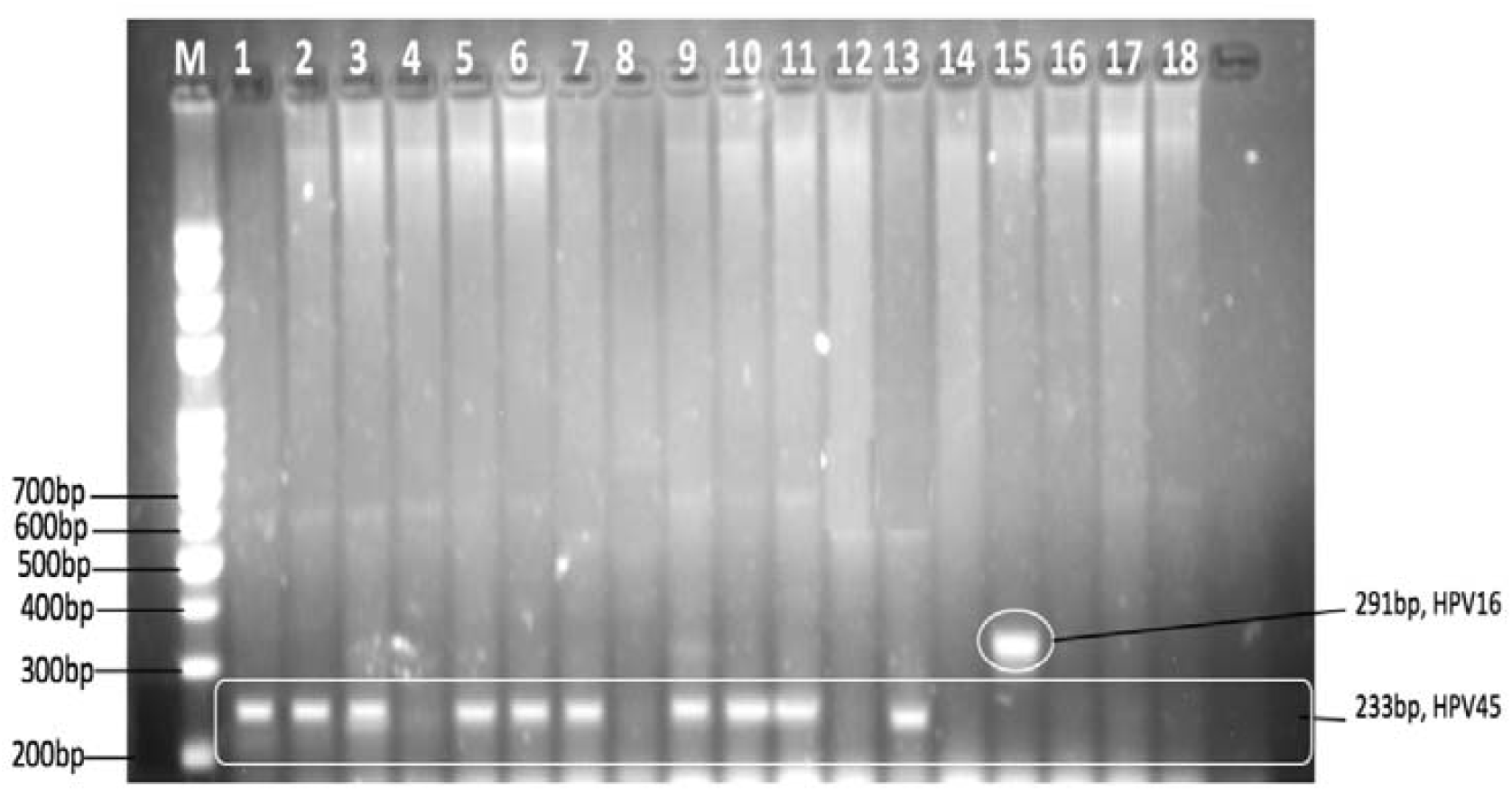
GEL A: Positive Polymerase Chain Reaction Bands With Corresponding positive HPV serotypes. **HPV DNA ANALYSIS**

**FIGURE 6:**
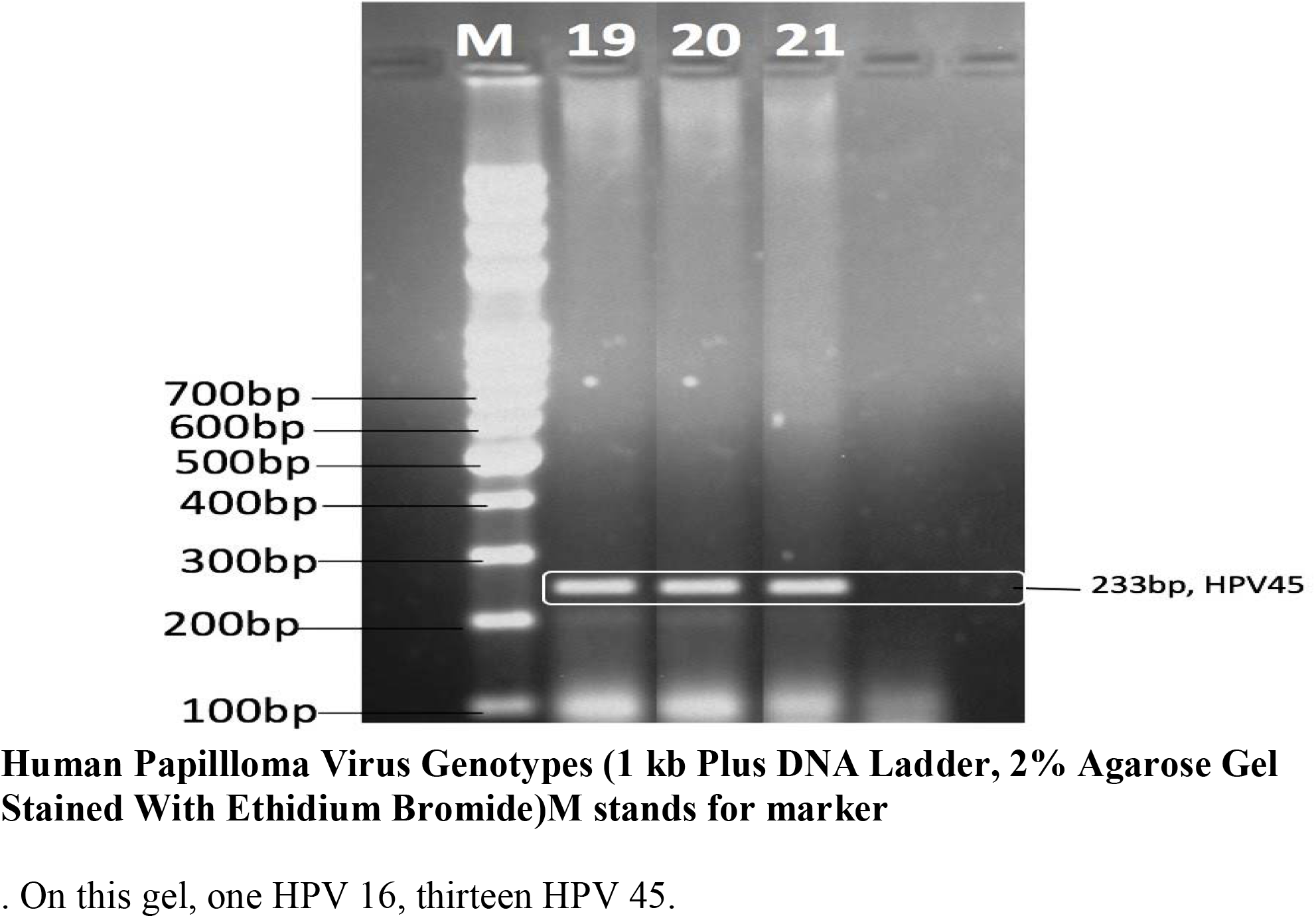
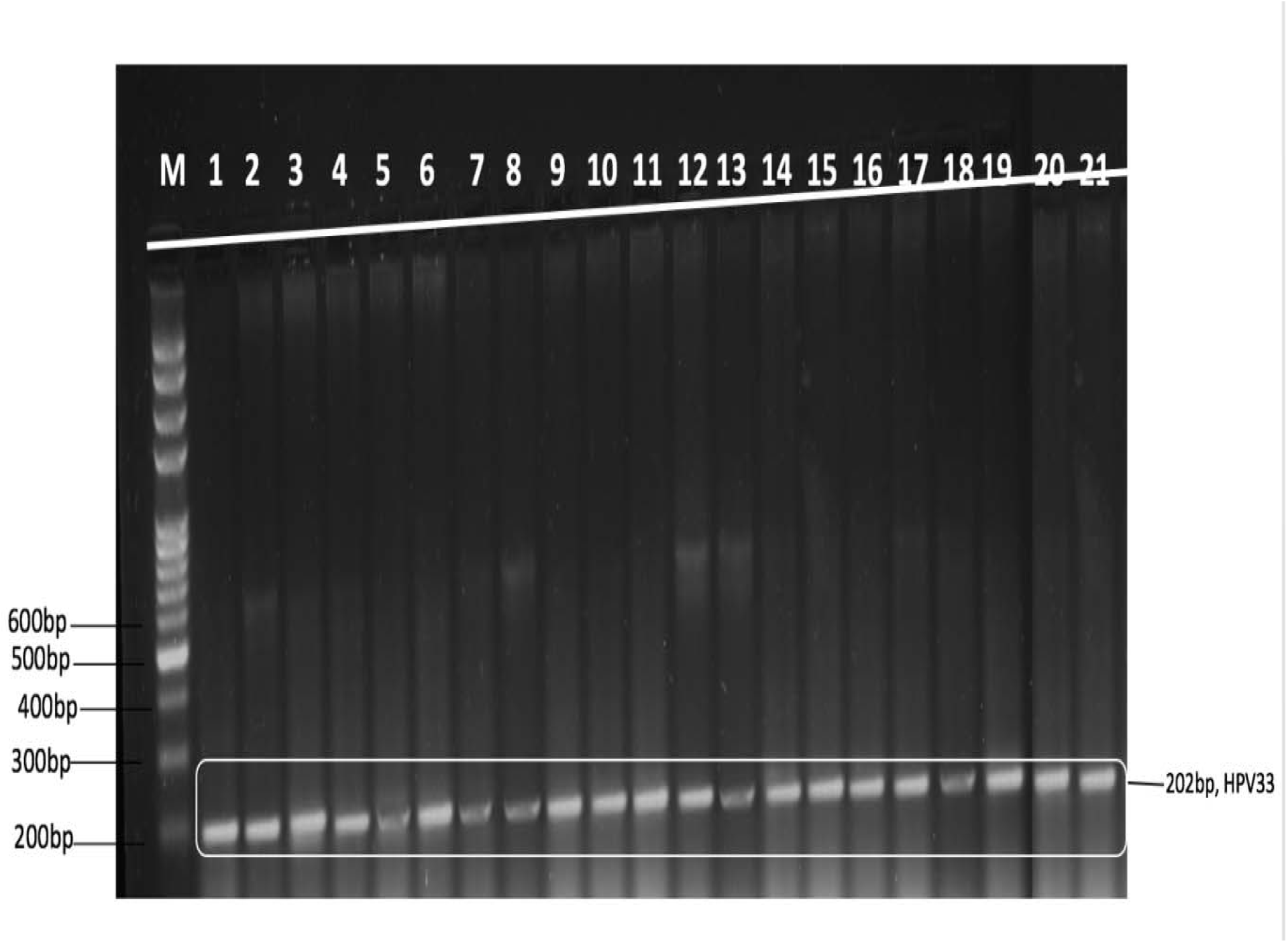
GEL B: Positive Polymerase Chain Reaction Bands With Corresponding. On this gel, all the samples contains HPV, DNA 33.

### Category of squamous intraepithelial lesion

Table 3 shows the various categories of squamous intraepithelial lesion. The only serotype found among study participants were HPV 16, 33 and 45. The only person with HPV 16 had high grade squamous intraepithelial lesion. Among those with HPV 33, 13(61.9%) were low grade while 8(38.1%) were high grade. Among those with HPV 45, 8(61.5%) were low grade while 5(38.5%) were high grade.

**Table 5:** Category of squamous intraepithelial lesion.

| **HPV serotype** | **Category of squamous intraepithelial lesion** |  |
| --- | --- | --- |
|  | **Low grade** | **High grade** |
| 16 | - | 1(4.7) |
| 18 | - | - |
| 33 | 13(61.9) | 8(38.1) |
| 45 | 8(61.5%) | 5(38.5) |

### Human Papillloma Virus Genotypes (1 kb Plus DNA Ladder, 2% Agarose Gel Stained With Ethidium Bromide)M stands for marker

. On this gel, one HPV 16, thirteen HPV 45.

On this gel, all the samples contains HPV, DNA 33.

## DISCUSSION

### Discussion/Conclusion

The Prevalence rate of high risk Human Papilloma virus in this study was 9.9%. The prevalence rate is higher when compared with previous study of 3% reported in a study done in Calabar in 2016 by Ago et al on prevalence of abnormal cervical cytology among postnatal clinic attendees at the University of Calabar Teaching Hospital, Nigeria [25]. The difference may be due to the different sample population. The previous study was a hospital based study where 100 women were recruited at antenatal during pregnancy and post natal ward after delivery while this study focused on non pregnant sexually active females aged 18-65years recruited from the three provinces of Qua, Efut and Efik Provinces.

The study done in Abuja by Modibbo et al on randomised trial evaluating self sampling for HPV DNA based test for cervical cancer screening in Nigeria showed a prevalence of 10%.[26]..The prevalence of hrHPV infection in this study was similar to other findings from previous studies of women who presented for cervical screening at hospitals in Abuja, Nigeria [27], [28].

This is much higher than the prevalence found in our study, probably because it was a community based study.

Studies done using Polymerase chain reaction (PCR)-based assays revealed higher HPV prevalence. A meta-analysis study done in Nigeria by <u>Kabuga</u> A.I et al showed overall prevalence of cervical papillomavirus infection was 42% in the general population [29].

However,23.7%in Mozambique [30],31% was seen in Harare, Zimbambwe[31], 60.3% among Colombian women [32], 65.0% in Gauteng[33], and 68.2% in the Western Cape provinces [34].

The higher prevalence in PCR-based studies may be defined by the different sensitivity of the test [30] - [34].

Out of 304 participants, 274(90%) were Negative for Squamous Intraepithelial lesion, out of this group 132(43.4%) had evidence of infection while 142(46.7%) did not show any evidence of infection. This is consistent with the study doneinCÃ’te d’ IvoirebyOuattara A. et al showed of the 339 women who took part in the study, 318 (93.8%) of 339 are negative visual inspection with acetic acid and Lugol [35].

Out of 30(9.9%) participants who were positive for Cervical Intraepithelial Lesions, 19 (63.3%) had low grade squamous intraepithelial Lesion (LSIL) and 11(36.6%) had high grade squamous intraepitheliallesion (HSIL). Out of these 30 participants, only 21 came for HPV DNA test. Of the 21 participants, 13(61.9%) were diagnosed with LSIL while the remaining 8(38.1%) had HSIL. Results showed that the highest number of abnormal smears were of low-grade squamous intraepithelial lesion (LSIL). This is similar to the results obtained by Obaseki and Nwafor, who reported that LSIL accounted for the highest percentage (66.27%) of abnormalities seen in cervical smears. [36], and also comparable with study done by Kolawole et al on molecular detection of Human Papillomavirus from abnormal cervical cytology of women attending a tertiary health facility in Ido-Ekiti, southwest Nigeria where Low-grade squamous intraepithelial lesions (LSIL) accounted for the majority of abnormal cytology, in 50% of the total abnormal smears. High-grade squamous intraepithelial lesions (HSIL) were 28.6%, while atypical squamous cells of undetermined significance (ASCUS) were 21.4%[37].The DNA typing was effectively used to verify the presence of HPV in the abnormal cytology.

In this study, HPV DNA was observed in all the 21 participants with abnormal cytology who had HPV DNA test. This is in line with study done by Kolawole et al in Ido-Ekiti south West of Nigeria where all the abnormal cervical smears contain HPV DNA [37].

The frequency of hr-HPV in abnormal cervical cytology had the strong relationship between cervical cancer, hrHPV DNA positivity and abnormal cervical cytology.

The prevailing high risk HPV serotypes seen among study participants were HPV 33 which occurred in 100%, followed by HPV 45 and HPV 16which occurred in 61.9% and 4.8% respectively.This is consistent with VIVIANEstudy, where the HPV33 and HPV16 were found to be associated with the highest risk of Cervical Intraepithelial Neoplasia (CIN) development, followed by HPV18, HPV31, and HPV45[12].

Even though, HPV 16/18 infections accounted for majority of the disease globally, the presence of HPV 16 to invasive cervical cancer from Sub-Saharan Africa, specifically West Africa is among the lowest worldwide [38].

In this study,66.67%(14) out of 21 participants who came out for HPV DNA had co – infection namely HPV 33 with HPV 45 (61.9%) and HPV 33 with HPV 16 (4.8%)respectively. Furthermore, 33.33% has single infections mainly HPV 33. This is consistent with study done by Pamela B. M et al Libreville, in Gabon where multiple HPV infections were observed in 42.31 % of HPV-infected women [39].

## CONCLUSION

The prevalence of high risk HPV in this study was 9.9% and most common in older female with the serotype 33 being the commonest. The highest number of abnormal smears were of low grade squamous intraepithelial lesion. HPV DNA was observed in all the abnormal cytology subjected for HPV DNA test.

The prevailing high risk HPV serotypes seen among study participants were HPV 33 which occurred in 100% followed by HPV 45 which was found in 61.9%,then HPV 16 which was seen in 4.76% participants. Among those with HPV 33,13(61.9%) were of low grade while 8(38.1%) were of high grade. Among those with HPV45, 8(38.1%) were low grade while 5(23.8%) were high grade. The only participant with HPV16 had high grade squamous intraepithelial lesion and constituent 4.76%.

In this study,66.67%(14) out of 21 participants had HPV DNA co – infection.

## Data Availability

All data produced in the present work are contained in the manuscript

## ACKNOWLEDGEMENTS

We would want to thank the Former and Present Chief Medical Directors of the University of Calabar Teaching Hospital, Professor AganThomos and Professor IkpemeIkpeme, respectively, the former and present Chairman Medical Advisory Committee of the hospital Professor NgimOgbu and Dr Michael Eyong for their immense support during this study. Our special thanks also go to both the Former and Present Heads of department of Pathology, Acting laboratory Directors, Department of Pathology University of Calabar Teaching Hospital; Dr Ebughe Godwin, Dr OmotosoAyodele, Dr. NwachukwuTobechukwu and Dr Ernest Neomi, respectively. Also to Professor AtimUdoh, Head of Department of Obstetrics and Gyneacology, University of Calabar Teaching Hospital, Mr Enya Jacob Afen for their cooperation and Mrs. Chigozie Augustina Udoka who stained all the cytological slide during the study.

