## Supplementary material for "HUMAN PAPILLOMAVIRUS SEROTYPES IN ABNORMAL CERVICAL SMEARS IN CALABAR CROSS RIVER STATE A CROSS SECTIONAL STUDY": TABLES

**Table 1: Age characteristics of study participants (N=304)**

| Variables | Frequency | Percentage (%) |
| --- | --- | --- |
| Age group(years) |  |  |
| 18-25 | 15 | 4.93 |
| 26-35 | 91 | 29.9 |
| 36-45 | 106 | 34.9 |
| 46-55 | 65 | 21.4 |
| 56-65 | 27 | 8.9 |
| Mean age ± SD* | 40.5 ± 10.2 |  |

SD*=Standard Deviation

**Table 2: Association between age characteristics and high risk HPV serotype predominance among study participants**

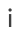

|  | **High risk HPV serotype** | | |  |  |
| --- | --- | --- | --- | --- | --- |
| **Variables** | **Yes**  **n(%)** | **No**  **n(%)** | **Total** | **Chi Square Test** | **P-value** |
| **Age group(years)** |  |  |  |  |  |
| 18-25 | 0(0.0) | 15(4.9) | 15(4.9) | FET;19.721 | <0.001* |
| 26-35 | 2(0.7) | 89(29.3) | 91(29.9) |  |  |
| 35-45 | 9(3.0) | 97(31.9) | 106(34.9) |  |  |
| 46-55 | 12(3.9) | 53(17.4) | 65(21.4) |  |  |
| 56-65 | 7(2.3) | 20(6.6) | 27(8.9) |  |  |
| ***TOTAL*** | ***30(9.9)*** | ***274(90.1)*** | ***304(100.0)*** |  |  |

The table above shows the aged range of the study between aged 18 to 65years and their aged intervals. Out of 304 (100), 30(9.9) were positive for abnormal cervical smears(either high grade or low grade squamous intraepithelial lesions) .The results were subjected to high risk HPV serotypes test and all tested positive. A total of 274(90.1) results were negative for abnormal cervical smears (negative for squamous intraepithelial lesion) and the results were not subjected to HPV DNA test.

**High risk HPV positive women**

Figure 2 below is a bar chart showing HPV serotype among correspondent with high

risk women. The most common serotype found among study participants was HPV

33 found among 100% (21), followed by HPV 45 found among 61.9%(13)then

HPV16 which was found among 4.7%(1) and HPV 18 was 0(none).

**Table 3: Category of squamous intraepithelial lesion**

|  | **Category of squamous intraepithelial lesion** | |
| --- | --- | --- |
| **HPV serotype** | **Low grade** | **High grade** |
| 16 | - | 1(4.7) |
| 18 | - | - |
| 33 | 13(61.9) | 8(38.1) |
| 45 | 8(61.5%) | 5(38.5) |
